# Predicting Axillary Lymph Node Metastasis in Early Breast Cancer Using Deep Learning on Primary Tumor Biopsy Slides

**DOI:** 10.1101/2021.10.10.21264721

**Authors:** Feng Xu, Chuang Zhu, Wenqi Tang, Ying Wang, Yu Zhang, Jie Li, Hongchuan Jiang, Zhongyue Shi, Jun Liu, Mulan Jin

**Affiliations:** Department of Breast Surgery, Beijing Chao-Yang Hospital, Beijing, 100020; School of Artificial Intelligence, Beijing University of Posts and Telecommunications, Beijing, 100876; Department of Pathology, Beijing Chao-Yang Hospital, Beijing, 100020

**Keywords:** Deep learning, Axillary lymph node metastasis, Breast cancer, Core-needle biopsy, Whole-slide images

## Abstract

**Objectives:** To develop and validate a deep learning (DL) based primary tumor biopsy signature for predicting axillary lymph node (ALN) metastasis preoperatively in early breast cancer (EBC) patients with clinically negative ALN.

**Methods:** A total of 1058 EBC patients with pathologically confirmed ALN status were enrolled from May 2010 to August 2020. A deep learning core-needle biopsy (DL-CNB) model was built on the attention based multiple instance learning (AMIL) framework to predict ALN status utilizing the deep learning features, which were extracted from the cancer areas of digitized whole-slide images (WSIs) of breast CNB specimens annotated by two pathologists. Accuracy, sensitivity, specificity, receiver operating characteristic (ROC) curves, and areas under the receiver operating characteristic curve (AUCs) were analyzed to evaluate our model.

**Results:** The best performing DL-CNB model with VGG16_BN as the feature extractor achieved an AUC of 0.816 (95% confidence interval (CI): 0.758, 0.865) in predicting positive ALN metastasis in the independent test cohort. Furthermore, our model incorporating the clinical data, which was called DL-CNB+C, yielded the best accuracy of 0.831 (95%CI: 0.775, 0.878), especially for patients younger than 50 years (AUC: 0.918, 95%CI: 0.825, 0.971). The interpretation of DL-CNB model showed that the top signatures most predictive of ALN metastasis were characterized by the nuclei features including density (*p*=0.015), circumference (*p*=0.009), circularity (*p*=0.010), and orientation (*p*=0.012).

**Conclusion:** Our study provides a novel deep learning-based biomarker on primary tumor CNB slides to predict the metastatic status of ALN preoperatively for patients with early breast cancer.

## Introduction

Breast cancer (BC) has become the greatest threat to women’s health worldwide (1). Clinically, identification of axillary lymph node (ALN) metastasis is important for evaluating the prognosis and guiding the treatment for BC patients (2). Sentinel lymph node biopsy (SLNB) has gradually replaced ALN dissection (ALND) to identify ALN status, especially for early breast cancer (EBC) patients with clinically negative lymph nodes. Although SLNB had the advantage of less invasiveness than ALND, SLNB still caused some complications such as lymphedema, axillary seroma, paraesthesia, and impaired shoulder function (3, 4). Moreover, SLNB has been considered a controversial procedure, owing to the availability of radionuclide tracers and the surgeon’s experience (5, 6). In fact, SLNB can be avoided if there are some reliable methods of preoperative prediction of ALN status for EBC patients.

Several studies intended to predict the ALN status by clinicopathological data and genetic testing score (7, 8). However, due to the relatively poor predictive values and high genetic testing costs, these methods are often limited. Recently, deep learning (DL) can perform high-throughput feature extraction on medical images and analyze the correlation between primary tumor features and ALN metastasis information. In a previous study, deep features extracted from conventional ultrasound and shear wave elastography (SWE) were used to predict ALN metastasis, presenting an area under the curve (AUC) of 0.796 in the test set (9). Nevertheless, SWE has not been integrated into routine clinical breast examinations in many hospitals. Another recent study demonstrated that the DL model based on diffusion weighted magnetic resonance imaging (DWI-MRI) database of 172 patients achieved an AUC of 0.852 for preoperative prediction of ALN metastasis (10), but the small sample size enrolled could not be representative.

Currently, DL has enabled rapid advances in computational pathology (11, 12). For example, DL methods have been applied to segment and classify glomeruli with different staining and various pathologic changes, thus achieving the automatic analysis of renal biopsies (13, 14); meanwhile, there has shown promise for colorectal cancer detection (15, 16) by DL based automatic colonoscopy tissue segmentation and classification; besides, the analysis of gastric carcinoma and precancerous status can also benefit from DL schemes (17, 18). More recently, for the ALN metastasis detection, it is reported that DL algorithms on digital lymph node pathology images achieved better diagnostic efficiency of ALN metastasis than pathologists (19, 20). In particular, the assistance of algorithm significantly increases the sensitivity of detection for ALN micro-metastases (21). In addition to diagnosis, several previous studies indicated that deep features based on whole slide images (WSIs) of postoperative tumor samples potentially improved the prediction performance of lymph node metastasis in a variety of cancers (20, 22). So far, there is no relevant research on preoperatively predicting ALN metastasis based on WSIs of primary breast cancer samples. In this study, we investigated a clinical data set of EBC patients treated by preoperative core needle biopsy (CNB) to determine whether DL models based on primary tumor biopsy slides could help to refine the prediction of ALN metastasis.

## Patients and Methods

### Patients

On approval by the Institutional Ethical Committees of Beijing Chaoyang Hospital affiliated to Capital Medical University, we retrospectively analyzed data from EBC patients with clinically negative ALN from May 2010 to August 2020. Written consent was obtained from all patients and their families.

The detailed inclusion criteria were as follows: (1) patients with CNB pathologically confirmed primary invasive breast cancer; (2) patients who underwent breast surgery with SLN biopsy or ALND; (3) baseline clinicopathological data including age, tumor size, tumor type, ER/PR/HER-2 status and the number of ALN metastasis were comprehensive; (4) complete concordance of molecular status was found between CNB and excision specimens; (5) no history of preoperative radiotherapy and chemotherapy; and (6) adequate volume of biopsy materials with three or more cores for each patient.

The exclusion criteria included the following: (1) patients with physically or imaging positive ALN; (2) missing postoperative pathology information; (3) missing wax blocks and hematoxylin-eosin (HE) slices; (4) low-quality HE slices or WSI images. The patient recruitment workflow was showed in Fig. 1.

**Figure 1.**
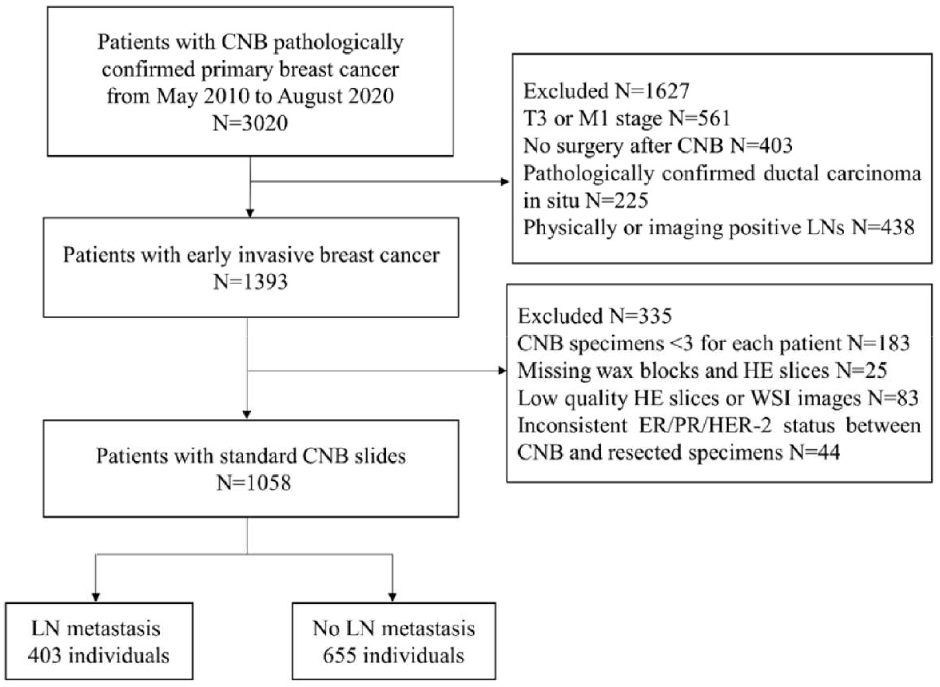
Patient recruitment workflow.

### Deep learning model development

To avoid the inter-observer heterogeneity, all available tumor regions in each CNB slide were examined and annotated by two independent and experienced pathologists blinded to all patient-related information. A WSI was classified into positive (N(+)) or negative (N0) using the proposed deep learning CNB (DL-CNB) model. Our DL-CNB model was constructed with the attention based multiple instance learning (MIL) approach (23). In MIL, each training sample was called a bag, which consisted of multiple instances (24-26) (each instance corresponds to an image patch of size 256×256 pixels). Different from the general fully-supervised problem where each sample had a label, only the label of bags was available in MIL, and the goal of MIL was to predict the bag label by considering all included instances comprehensively. The whole algorithm pipeline comprised the following five steps:

1. **Training data preparation (Fig. 2a)**. For each raw WSI, amounts of non-overlapping square patches were first cropped from the selected tumor regions. Then each WSI could be represented as a bag with *N* randomly selected patches. To increase the training samples, *M* bags were built for each WSI. All *M* bags were labeled as positive if the slide is an ALN metastasis case, and vice versa. Note that we could add the clinical information of the slide to all the *M* constructed bags to involve more useful information for predicting, and in this situation, the developed model was called DL-CNB+C.
2. **Feature extraction (left part of Fig. 2b)**. *N* feature vectors were extracted for the *N* image instances in each bag by using a convolutional neural network (CNN) model. The performances of AlexNet (27), VGG16 (28) with batch norm (VGG16_BN), ResNet50 (29), DenseNet121 (30), and Inception-v3 (31) were compared to find the best feature extractor. At this stage, the clinical data were also preprocessed for feature extraction. Concretely, the numerical properties in clinical data were standardizing by removing the mean and scaling to unit variance, thus eliminating the effect of data range and scale; furthermore, considering that there was not a natural ordinal relationship between different values of the category attributes, the categorical properties in clinical data were encoded as the one-hot vectors, which could express different values equally.
3. **MIL (right part of Fig. 2b)**. The extracted *N* feature vectors of image instances were first processed by the max-pooling (32-34) and reshaping, and then were passed to a two-layer fully connected (FC) layer. The *N* weight factors for the instances in the bag were thus obtained and then were further multiplied to the original feature vectors (23) for adaptively adjusting the effect of instance features. Finally, the weighted image feature vectors and the clinical features were fused by concatenation, due to the large difference of dimensions between image features and clinical features, the clinical features were copied 10 times for expansion. Then, the fused features were fed into the classifier, and the outputs and the ground truth labels were used to calculate the cross-entropy loss.
4. **Model training and testing**. We randomly divided the WSIs into training cohort and independent test cohort with the ratio of 4:1, and randomly selected 25% of the training cohort as the validation cohort. We used Adam optimizer with learning rate 1e-4 to update the model parameters, and weight decay 1e-3 for regularization. In the training phase, we used the cosine annealing warm restarts strategy to adjust the learning rate (35). In the testing phase, the ALN status is predicted by aggregating the model outputs of all bags from the same slide **(Fig. 2c)**. The deep learning models are available at: https://github.com/bupt-ai-cz/BALNMP.

**Figure 2.**
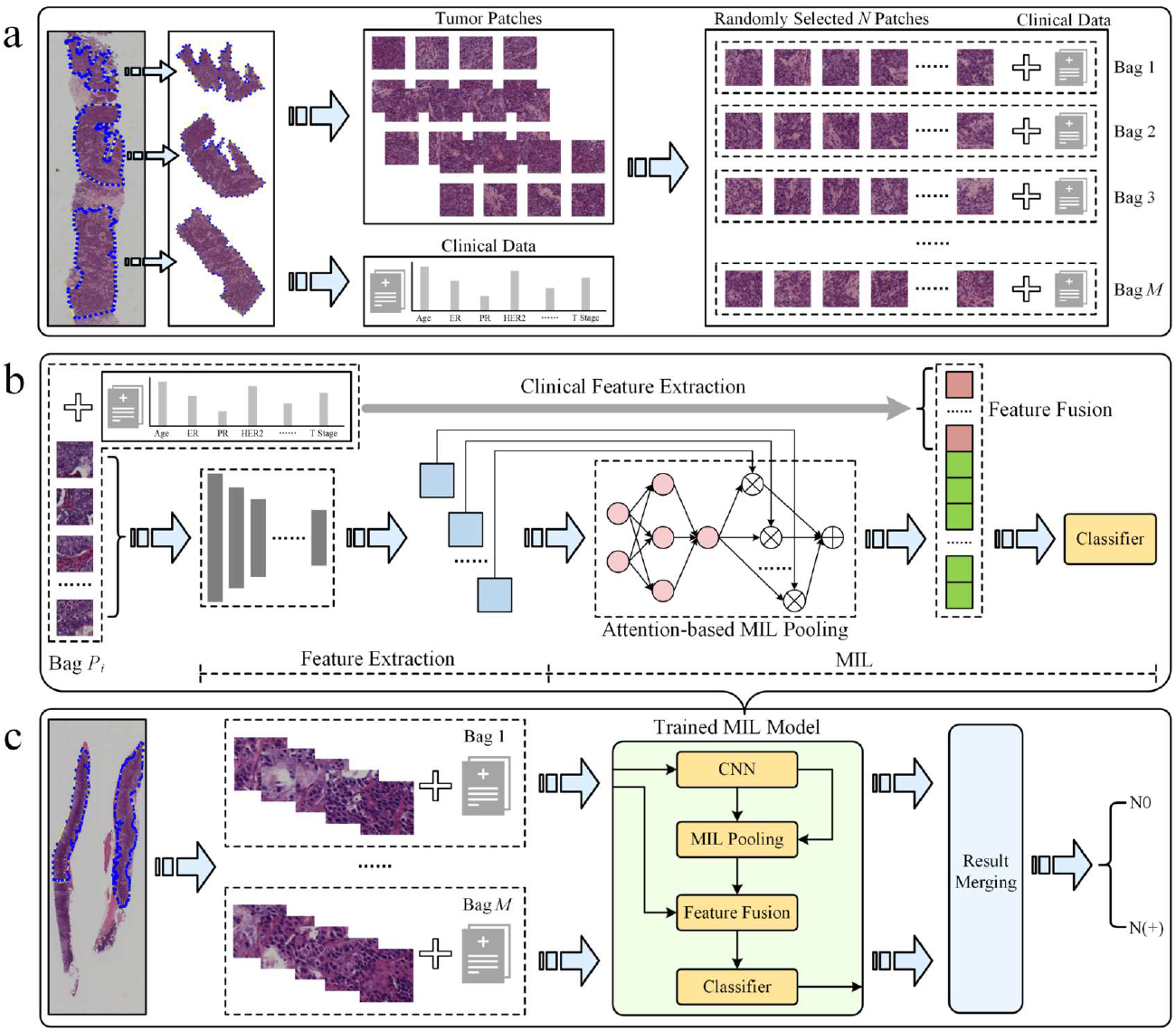
The overall pipeline of the DL-CNB+C model to predict ALN status between N0 and N(+). (a) Multiple training bags were built based on clinical data and the cropped patches from the selected tumor regions of each core needle biopsy (CNB) whole slide image (WSI). (b) DL-CNB+C model training process included two phases of feature extraction and multiple instance learning (MIL), and finally the weighted features fused with clinical features were used to predict classification probabilities and calculate the cross-entropy loss. (c) The predicted probabilities of each bag from a raw CNB WSI were merged to guide the final axillary lymph node (ALN) status classification between N0 and N(+).

### Visualization of salient regions from DL-CNB model

We visualized the important regions which were more associated with metastatic status. After the processing of attention-based MIL pooling, the weights of different patches can be obtained and the corresponding feature maps were then weighted together in the following FC layers to conduct ALN status prediction. With the attention weights, we created a heat map to visualize the important salient regions in each WSI.

### Interpretability of DL-CNB model with nuclei features

Interpretability of DL-CNB model with nuclei features was performed to study the contribution of different nuclei morphological characteristics in the prediction of lymph node metastasis (36, 37). Multiple specially designed nuclei features were firstly extracted for each WSI, and these features together formed a training bag. With the constructed feature bags, the proposed DL-CNB model was re-trained. The weights of different features (instances) can be obtained based on the attention-based MIL pooling, and thus the contribution of different features was yielded. The specific process was described in Fig. 3.

**Figure 3.**
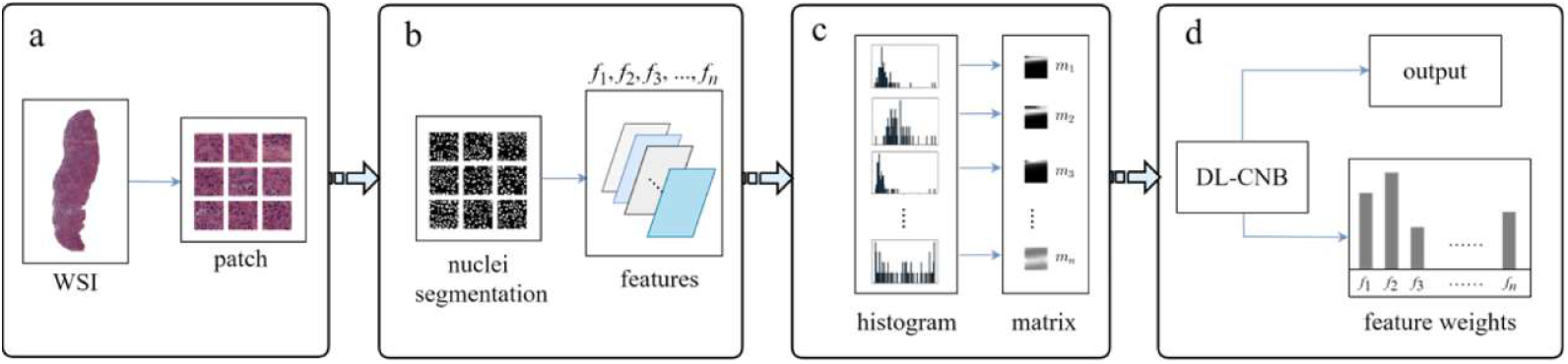
Overview on interpretability methods of DL-CNB model based on nuclei morphometric features. (a) The selected tumor regions of each WSI was cropped into patches. (b) For each patch, we processed nuclei segmentation (a weakly supervised segmentation framework was applied to obtain the nucleus), defined multiple nuclei morphometric features (such as major axis, minor axis, area, orientation, circumference, density, circularity and rectangularity, which are denoted as *f*_1_, *f*_2_, *f*_3_, …, *f*_n_) and extracted *n* feature parameters correspondingly. (c) All *n* kinds of feature parameters from a WSI were quantized into *n* distribution histograms and saved to *n* feature matrices (*m*_1_, *m*_2_, *m*_3_, …, *m*_n_). (d) The matrices from a WSI were considered as instances of a bag and served as the input of DL-CNB model; the re-trained DL-CNB model could generate scores of features (instances) in the bag, which represented the weight of each feature in pathological diagnosis.

### Statistical analysis

The logistic regression was used to predict ALN status by clinical data only model. The clinical difference of N0 and N(+) was compared by using the Mann-Whitney U test and chi-square test. The AUCs of different methods were compared by using Delong et al (38). The other measurements like accuracy (ACC), sensitivity (SENS), specificity (SPEC), positive predictive value (PPV), and negative predictive value (NPV) were also used to estimate the model performance. All the statistics were two-sided and a *p*-value less than 0.05 was considered statistically significant. All statistical analyses were performed by MedCalc software (V 19.6.1; 2020 MedCalc Software bvba, Mariakerke, Belgium), Python 3.7, and SPSS 24.0 (IBM, Armonk, NY).

## Results

### Clinical characteristics

A total of 1058 patients with early breast cancer were enrolled for analysis. Among them, 957 (90.5%) patients had invasive ductal carcinomas, and 101 (9.5%) patients had invasive lobular carcinomas. There were 840 patients in the training cohort and 218 patients in the independent test cohort after all WSIs were randomly divided by using N0 as the negative reference standard and others as the positive. The average patient age was 57.6 years (range, 26-90 years) for the training and validation sets, 56.7 years (range, 22-87 years) for the test set. The mean ultrasound tumor size was 2.23 cm (range, 0.5-4.5 cm). A total of 556 patients (52.6%) had T1 tumors, while 502 patients (47.4%) had T2 tumors. According to the results of SLNB or ALND, positive lymph nodes were found in 403 patients. Among them, 210 patients (52.1%) had one or two positive lymph nodes (N_+_(1-2)) and 193 patients (47.9%) had three or more positive lymph nodes (N_+_(≥3)). As shown in Table 1, there was no significant difference between the detailed characteristics of the training and independent test cohort (all *p* > 0.05).

**Table 1.**
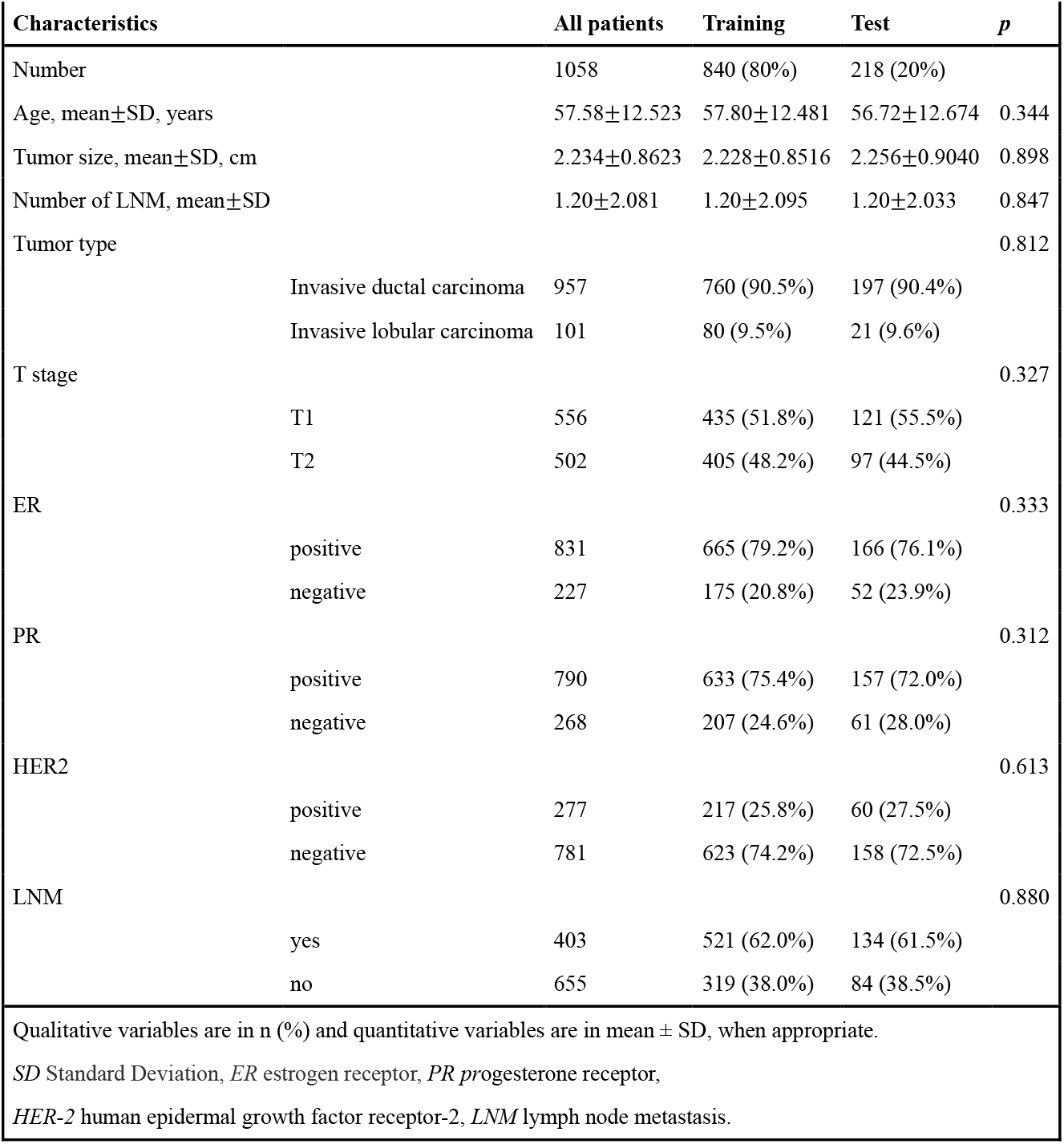
Patient and tumor characteristics.

### CNN model selection

The detailed results were summarized in supplemental Table 1. Based on the overall analysis, VGG16_BN model pre-trained on ImageNet (39) provided the best performance in the validation cohort and the independent test cohort (AUC: 0.808, 0.816), compared with AlexNet (AUC: 0.764, 0.780), ResNet50 (AUC: 0.644, 0.607), DenseNet121 (AUC: 0.714, 0.739), and Inception-v3 (AUC: 0.753, 0.762). Furthermore, considering other metrics, VGG16_BN achieved the best ACC, SPEC, and PPV in the independent test cohort. VGG16_BN consisted of (convolution layer, batch normalization layer, ReLU) as the basic block where ReLU played a role of activation function to provide the non-linear capability, and max-pooling layers were inserted between basic blocks for down-sampling, besides, there was an adaptive average pooling layer at the end of VGG16_BN for obtaining features with a fixed size. The details of VGG16_BN was described in supplemental Table 2.

### Predictive value of DL-CNB+C model between N0 and N(+)

In the training cohort, DL-CNB+C achieved the AUC of 0.878, while DL-CNB and classification by clinical data only model achieved AUCs of 0.901 and 0.661, respectively. And in the validation cohort, the DL-CNB+C model achieved the AUC of 0.823, which was higher than the AUC of 0.808 got by DL-CNB only and the AUC of 0.709 got by classification by clinical data.

In the independent test cohort, the DL-CNB+C model still achieved the highest AUC of 0.831 which was better than the AUC of DL-CNB only (AUC: 0.816, *p* = 0.453) and classification by clinical data only (AUC: 0.613, *p* < 0.0001). The ACC, SENS, and NPV of DL-CNB+C were also better than other methods. The detailed statistical results were summarized in Table 2 and its corresponding ROCs were shown in Fig. 4.

**Table 2.** The performance in prediction of ALN status (N0 vs. N(+)).

| Table 2 The performance in prediction of ALN status (N0 vs. N(+)). |  |  |  |  |  |  |  |
| --- | --- | --- | --- | --- | --- | --- | --- |
| Methods |  | AUC | ACC (%) | SENS (%) | SPEC (%) | PPV (%) | NPV (%) |
| Clinical data only | T | 0.661 [0.622, 0.698] | 64.13 [60.24, 67.88] | 64.58 [58.17, 70.63] | 63.85 [58.86, 68.62] | 52.36 [48.32, 56.38] | 74.55 [70.85, 77.92] |
|  | V | 0.709 [0.643, 0.770] | 67.62 [60.84, 73.90] | 65.82 [54.29, 76.13] | 68.70 [60.02, 76.52] | 55.91 [48.46, 63.11] | 76.92 [70.62, 82.22] |
|  | I-T | 0.613<sup>a, b</sup> [0.545, 0.678] | 61.93 [55.12, 68.40] | 50.00 [38.89, 61.11] | 69.40 [60.86, 77.07] | 50.60 [42.34, 58.83] | 68.89 [63.49, 73.82] |
| DL-CNB model | T | 0.901 [0.875, 0.923] | 80.32 [76.99, 83.35] | 94.17 [90.41, 96.77] | 71.79 [67.05, 76.21] | 67.26 [63.61, 70.71] | 95.24 [92.30, 97.09] |
|  | V | 0.808 [0.748, 0.859] | 72.86 [66.31, 78.75] | 77.22 [66.40, 85.90] | 70.23 [61.62, 77.90] | 61.00 [53.95, 67.62] | 83.64 [77.04, 88.62] |
|  | I-T | 0.816<sup>c</sup> [0.758, 0.865] | 74.77 [68.46, 80.39] | 80.95 [70.92, 88.70] | 70.90 [62.43, 78.42] | 63.55 [56.76, 69.84] | 85.59 [79.04, 90.34] |
| DL-CNB+C model | T | 0.878 [0.622, 0.698] | 76.51 [73.00, 79.77] | 93.33 [89.40, 96.14] | 66.15 [61.22, 70.84] | 62.92 [59.53, 66.19] | 94.16 [90.90, 96.30] |
|  | V | 0.823 [0.765, 0.872] | 75.71 [69.34, 81.35] | 74.68 [63.64, 83.80] | 76.34 [68.12, 83.32] | 65.56 [57.69, 72.65] | 83.33 [77.19, 88.08] |
|  | I-T | 0.831 [0.775, 0.878] | 75.69 [69.44, 81.23] | 89.29 [80.63, 94.98] | 67.16 [58.53, 75.03] | 63.03 [56.96, 68.71] | 90.91 [84.21, 94.94] |
| 95% confidence intervals are included in brackets.<br>*AUC* area under the receiver operating characteristic curve, *ACC* accuracy, *SENS* sensitivity, *SPEC* specificity, *PPV* positive predict value, *NPV* negative predict value.<br>*T* training cohort (n = 630), *V* validation cohort (n = 210), *I-T* independent test cohort (n = 218).<br><sup>a</sup>Indicates $p < 0.0001$, Delong et al. in comparison with DL-CNB model in independent test cohort.<br><sup>b</sup>Indicates $p < 0.0001$, Delong et al. in comparison with DL-CNB+C model in independent test cohort.<br><sup>c</sup>Indicates $p = 0.4532$, Delong et al. in comparison with DL-CNB+C model in independent test cohort. | | | | | | | |

**Figure 4.**
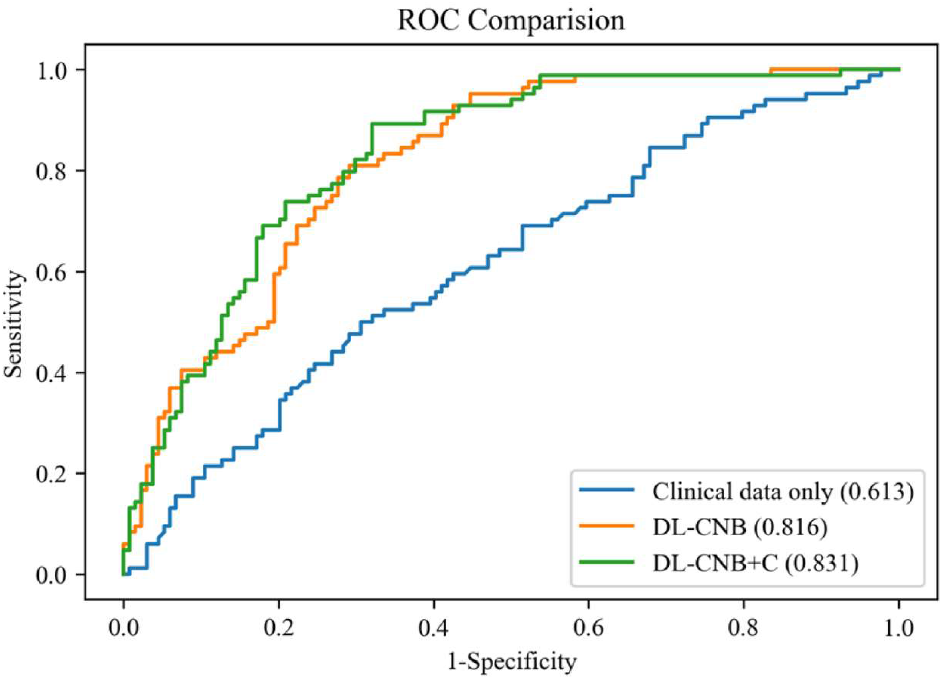
Comparison of receiver operating characteristic (ROC) curves between different models for predicting disease-free axilla (N0) and heavy metastatic burden of axillary disease (N(+)). Numbers in parentheses are AUCs.

We further divided N(+) into low metastatic potential (N_+_(1-2)) and high metastatic potential (N_+_(≥3)) according to the number of ALN metastasis. Adopting N0 as the negative reference standard, the combined model showed better discriminating ability between N0 and N_+_(1-2) (AUC: 0.878), between N0 and N_+_(≥3) (AUC: 0.838).

The detailed statistical results were summarized in supplemental Table 3 and Table 4, and the corresponding ROCs were shown in supplemental Fig. 1 and Fig. 2.

### Predictive value of DL-CNB+C model among N0, N_+_(1-2) and N_+_(≥3)

The overall AUC of multi-classification in the independent test cohort based on DL-CNB +C model was 0.791, there existed the highest precision and recall of 0.747 and 0.947 respectively in N0, there existed the precision and recall of 0.556 and 0.400 in N_+_(1-2), and there existed the precision and recall of 0.375 and 0.162 in N_+_(≥3). The confusion matrix under the classification threshold of 0.5 was shown in Fig. 5. According to the results, the model performed well in differentiating the N0 group while showed poor diagnostic efficacy in the other two groups.

**Figure 5.**
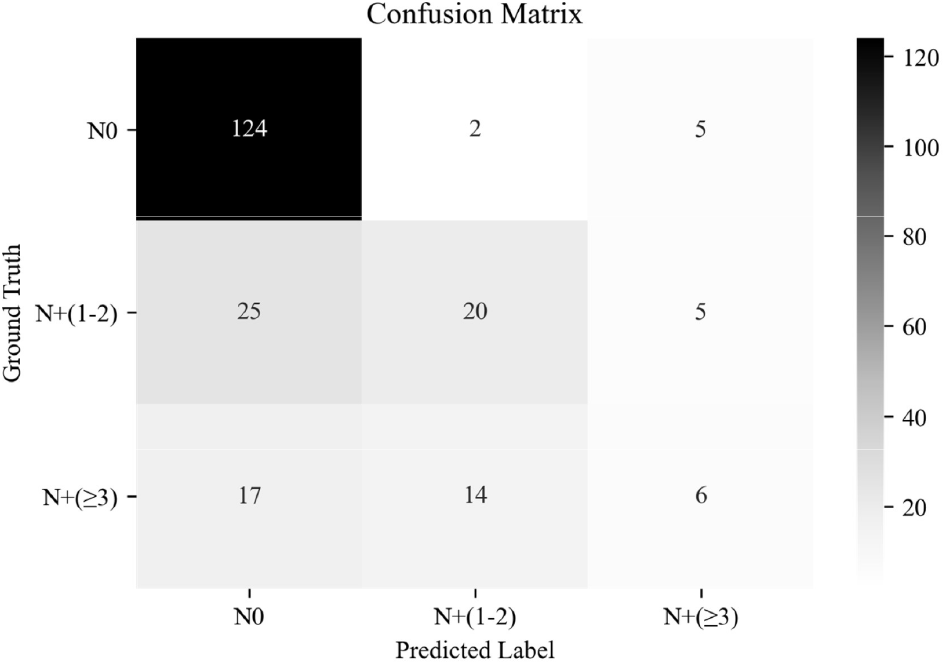
The confusion matrix of predicting ALN status between disease-free axilla (N0), low metastatic burden of axillary disease (N+(1-2)) and heavy metastatic burden of axillary disease (N+(≥3)).

### Subgroup analysis of DL-CNB+C model

Furthermore, we analyzed the measurement results of the different subgroups in the independent test cohort of predicting ALN status between N0 and N(+) by the DL-CNB+C model. The detailed statistical results were summarized in supplemental Table 5. In the independent test cohort, compared with the AUC of 0.794 (95%CI: 0.720, 0.855) in the subgroup of age > 50, there existed better performance in the subgroup of age ≤ 50 with the AUC of 0.918 (95%CI: 0.825, 0.971, *p* = 0.015). There were no significant differences regarding other subgroups of ER(+) vs. ER(-) (*p* = 0.125), PR(+) vs. PR(-) (*p* = 0.659), HER-2(+) vs. HER-2(-) (*p* = 0.524), T1 vs. T2 stage (*p* = 0.743) between N0 and N(+).

### Interpretability of DL-CNB model

To investigate the interpretability of the DL-CNB, we conducted two studies for digging the correlation factors of ALN status prediction. In the first study, we adopted the attention-based MIL pooling to find the important regions that contributing to the prediction. The heat map in Fig. 6a highlights the red patches as the important regions. Although the obtained important areas can provide some clues to the diagnosis of DL-CNB model, it is not clear that the model makes decisions based on what features of the tumor area.

**Figure 6.**
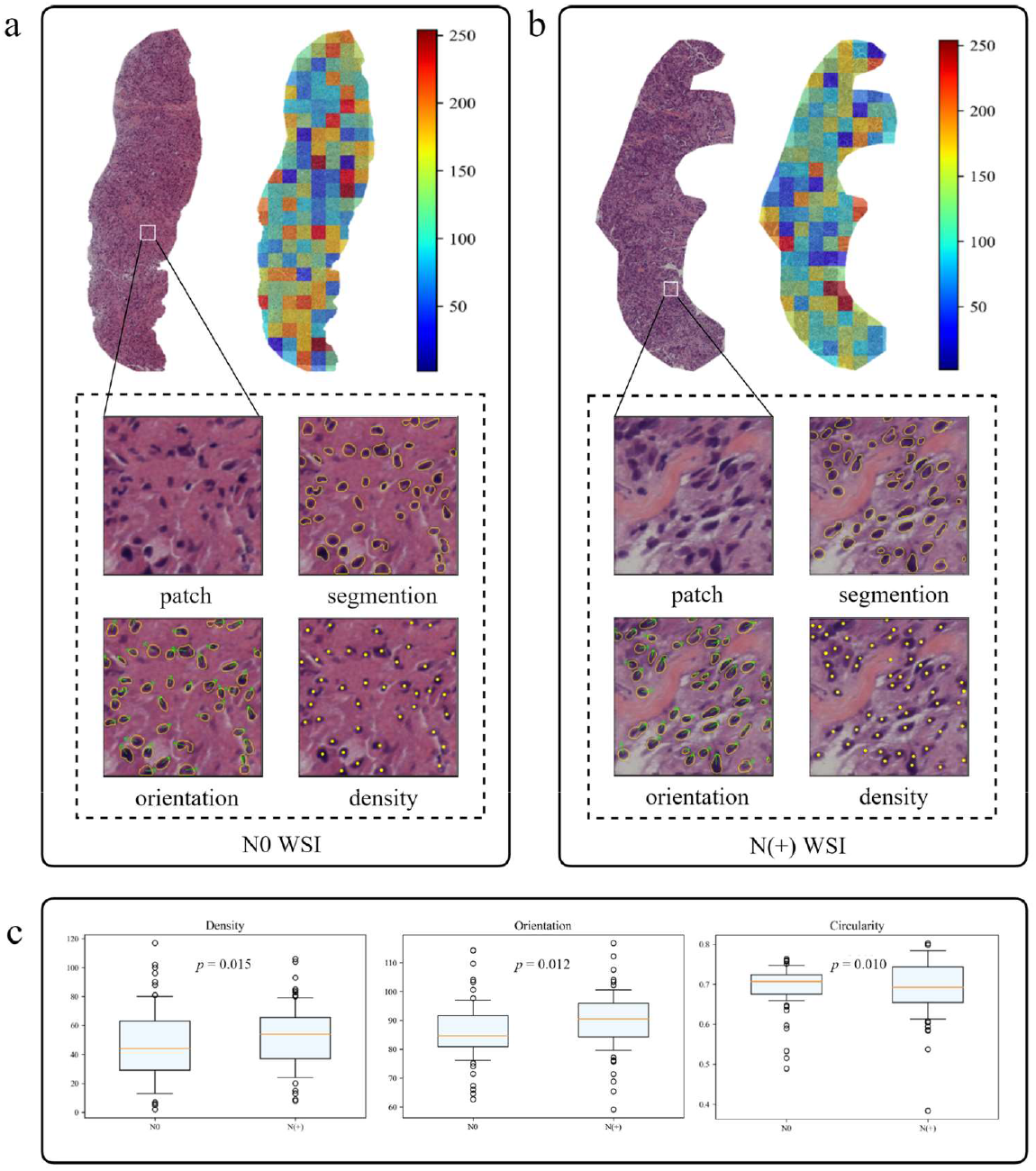
The interpretability of the DL-CNB model of two patients. In (a) and (b), we presented the heat maps and nuclear segmentation from core needle biopsy (CNB) whole slide images (WSIs) of the N0 and the N(+) separately, and the red regions showed greater contribution to the final classification. (c) showed the statistical analysis of three nuclear characteristics most relevant to diagnosis of all patients.

In the second study, we specially designed and extracted multiple nuclei features for each WSI. The weights of different features were then obtained based on the same attention-based MIL pooling in our DL-CNB. The weights highlighted the nuclei features that were most relevant to the ALN status prediction of each WSI. We found that the WSI of N(+) group had higher nuclear density (*p*=0.015) and orientation (*p*=0.012) but lower circumference (*p*=0.009), circularity (*p*=0.010) and area (*p*=0.024) compared with N0 group (Fig. 6b and 6c). There were no significant differences in other nuclei features including major axis (*p*=0.083), minor axis (*p*=0.065), rectangularity (*p*=0.149) between N0 and N(+).

## Discussion

In most previous studies, DL signatures of ALN metastases were based on medical images such as ultrasound, CT, and MRI images (10, 40, 41). However, since many patients had undergone CNB at the time of imaging examination, and the reactive changes such as needle path in the tumor would result in the predictive inaccuracy of imaging information. This study focused on preoperative CNB WSI, which also played an important role in breast cancer management and has been increasingly performed in clinical practice. Preoperative CNB can provide not only the histopathological diagnosis of breast cancer but also the molecular status including ER/PR/HER-2 status, which is associated with ALN metastasis(42). Otherwise, the morphological features of tumor cells can be visualized on CNB WSI. Therefore, primary tumor biopsy WSI as a complementary imaging tool has the potential for ALN metastasis prediction. To the best of our knowledge, this is the first study to apply the deep learning-based histopathological features extracted from primary tumor WSIs for ALN prediction analysis.

Here, the best-performing DL-CNB model yielded satisfactory predictions with an AUC of 0.816, a SENS of 81.0%, and a SPEC of 70.9% on the test set, which had superior predictive capability compared with clinical data alone. Furthermore, unlike other combined models incorporating clinical data (7, 9), the DL-CNB+C model slightly improved the ACC to 0.831, which showed that our results were mainly derived from the contribution of DL-CNB model. In addition, during the subgroup analysis stratified by patient’s age, our DL-CNB+C model achieved an AUC of 0.918 for patients younger than 50 years, indicating that age was the critical factor in predicting ALN status. Regarding the number of ALN metastasis, the DL-CNB+C model showed better discriminating ability between N0 and N_+_(1-2), between N0 and N_+_(≥3). However, the unfavorable discriminating ability was found between N_+_(1-2) and N_+_(≥3). This was consistent with the study of Zheng et al. (9) who also reported the poor efficacy between N_+_(1-2) and N_+_(≥3) utilizing the DL radiomics model. In the future, further exploration of ALN staging prediction is needed.

Indeed, computer-assisted histopathological analysis can provide a more practical and objective output (43). For example, different molecular subtypes (44) and Oncotype DX risk score (45) occurring in breast cancer could be directly predicted from the HE slides. On one hand, our DL model can provide significant information for risk stratification and axillary staging, thereby avoiding axillary surgery and reducing the complication and hospitalization costs. On the other hand, our results also highlight the development of algorithms based on artificial intelligence, which will reduce the labor intensity of pathologists. Similar approaches may be used to the pathology of other organs.

In our study, we are first to quantitatively assess the role of nuclear disorder in predicting ALN metastasis in breast cancer. Our finding is consistent with several recent studies that demonstrate the powerful predictive effect of nuclear disorder on patient survival (46, 47). Interestingly, the top predictive signatures that distinguished N0 from N(+) were characterized by the nuclei features including density, circumference, circularity, and orientation. We found that the WSI of N(+) had higher nuclear density and polarity but lower circularity, which was understandable since in the tumors with ALN metastasis, tumor cells became poorly differentiated as a result of rapid cell growth, encouraging the nuclei in these structures to form highly clustered and consistently metastatic patterns. Our results showed that nuanced patterns of nuclei density and orientation of tumor cells are important determinants of ALN metastasis.

There are some limitations in our study. First, the selection of regions of interest within each CNB slide required pathologist guidance. Future studies will explore more advanced methods for automatic segmentation of tumor regions. Secondly, this is a retrospective study, prospective validation of our model in a large multicenter cohort of early breast cancer patients is necessary to assess the clinical applicability of the biomarker. Thirdly, recent evidence indicated that a set of features related to tumor-infiltrating lymphocytes (TILs) was found to be associated with positive LNs in bladder cancer (22). However, due to few TILs on breast CNB slides, we only selected sufficient tumor cells for the identification of salient regions rather than whole slides. Finally, we only chose HE stained images of CNB samples. The clinical utility of immunochemical stained images remains to be established as an interesting attempt.

## Conclusion

In brief, we demonstrated that a novel deep learning-based biomarker on primary tumor CNB slides predicted ALN metastasis preoperatively for EBC patients with clinically negative ALN, especially for younger patients. Our methods could help to avoid unnecessary axillary surgery based on the widely collected HE stained histopathology slides, thereby contributing to precision oncology treatment.

## Data Availability

All data produced in the present study are available upon reasonable request to the authors

## Ethics approval and consent to participate

This study was approved by the institutional review board of Beijing Chao-Yang Hospital, Capital Medical University. All patients provided written informed consent.

## Authors’ contributions

FX, CZ, JL, YW, and MJ designed the study. CZ, WT, YZ, JL trained the model. FX, YW, ZS, JL, and HJ collected the data. FX, WT, YZ, CZ, YW, MJ and JL analyzed and interpreted the data. FX, CZ, WT, YZ, and MJ prepared the manuscript. All authors read and approved the final manuscript.

## Data Availability

Excel files containing raw data included in the main figures and tables can be found in the Source Data File in the article. All other data are available in the Article and Supplementary Information. All other data including the imaging data can be provided upon reasonable request to the corresponding author.

## Supplemental Materials

**Table 1.** The performance comparison of different base models in prediction of ALN status (N0 vs. N(+)).

|  |  |  |  |  |  |  |  |
| --- | --- | --- | --- | --- | --- | --- | --- |
| Base models |  | AUC | ACC (%) | SENS (%) | SPEC (%) | PPV (%) | NPV (%) |
| AlexNet | T | 0.909 [0.884, 0.930] | 82.70 [79.51, 85.57] | 88.33 [83.58, 92.11] | 79.23 [74.86, 83.15] | 72.35 [68.20, 76.16] | 91.69 [88.59, 94.01] |
|  | V | 0.764 [0.700, 0.819] | 65.71 [58.87, 72.11] | 89.87 [81.02, 95.53] | 51.15 [42.26, 59.97] | 52.59 [47.84, 57.30] | 89.33 [80.96, 94.28] |
|  | I-T | 0.780 [0.719, 0.833] | 73.39 [67.01, 79.13] | 83.33 [73.62, 90.58] | 67.16 [58.53, 75.03] | 61.40 [55.08, 67.36] | 86.54 [79.71, 91.32] |
| ResNet50 | T | 0.912 [0.887, 0.933] | 85.71 [82.74, 88.35] | 85.83 [80.77, 89.99] | 85.64 [81.76, 88.97] | 78.63 [74.17, 82.50] | 90.76 [87.77, 93.08] |
|  | V | 0.644 [0.575, 0.709] | 59.52 [52.55, 66.22] | 70.89 [59.58, 80.57] | 52.67 [43.77, 61.45] | 47.46 [41.80, 53.19] | 75.00 [67.22, 81.44] |
|  | I-T | 0.607 [0.539, 0.673] | 58.72 [51.87, 65.32] | 66.67 [55.54, 76.58] | 53.73 [44.92, 62.38] | 47.46 [41.61, 53.37] | 72.00 [64.65, 78.33] |
| DenseNet121 | T | 0.967 [0.949, 0.979] | 89.84 [87.21, 92.09] | 95.83 [92.47, 97.98] | 86.15 [82.32, 89.42] | 80.99 [76.85, 84.53] | 97.11 [94.82, 98.41] |
|  | V | 0.714 [0.648, 0.774] | 68.57 [61.82, 74.79] | 73.42 [62.28, 82.73] | 65.65 [56.85, 73.72] | 56.31 [49.56, 62.84] | 80.37 [73.56, 85.77] |
|  | I-T | 0.739 [0.675, 0.796] | 69.27 [62.68, 75.32] | 85.71 [76.38, 92.39] | 58.96 [50.13, 67.37] | 56.69 [51.21, 62.02] | 86.81 [79.28, 91.89] |
| Inception-v3 | T | 0.968 [0.951, 0.980] | 91.75 [89.32, 93.77] | 95.42 [91.95, 97.69] | 89.49 [86.01, 92.35] | 84.81 [80.68, 88.20] | 96.94 [94.68, 98.26] |
|  | V | 0.753 [0.689, 0.810] | 70.48 [63.81, 76.55] | 67.09 [55.60, 77.25] | 72.52 [64.04, 79.95] | 59.55 [51.71, 66.93] | 78.51 [72.39, 83.59] |
|  | I-T | 0.762 [0.700, 0.817] | 71.10 [64.59, 77.02] | 85.71 [76.38, 92.39] | 61.94 [53.16, 70.18] | 58.54 [52.79, 64.06] | 87.37 [80.12, 92.23] |
| VGG16_BN | T | 0.901 [0.875, 0.923] | 80.32 [76.99, 83.35] | 94.17 [90.41, 96.77] | 71.79 [67.05, 76.21] | 67.26 [63.61, 70.71] | 95.24 [92.30, 97.09] |
|  | V | 0.808 [0.748, 0.859] | 72.86 [66.31, 78.75] | 77.22 [66.40, 85.90] | 70.23 [61.62, 77.90] | 61.00 [53.95, 67.62] | 83.64 [77.04, 88.62] |
|  | I-T | 0.816 [0.758, 0.865] | 74.77 [68.46, 80.39] | 80.95 [70.92, 88.70] | 70.90 [62.43, 78.42] | 63.55 [56.76, 69.84] | 85.59 [79.04, 90.34] |
| VGG16_BN+C | T | 0.878 [0.622, 0.698] | 76.51 [73.00, 79.77] | 93.33 [89.40, 96.14] | 66.15 [61.22, 70.84] | 62.92 [59.53, 66.19] | 94.16 [90.90, 96.30] |
|  | V | 0.823 [0.765, 0.872] | 75.71 [69.34, 81.35] | 74.68 [63.64, 83.80] | 76.34 [68.12, 83.32] | 65.56 [57.69, 72.65] | 83.33 [77.19, 88.08] |
|  | I-T | 0.831 [0.775, 0.878] | 75.69 [69.44, 81.23] | 89.29 [80.63, 94.98] | 67.16 [58.53, 75.03] | 63.03 [56.96, 68.71] | 90.91 [84.21, 94.94] |
| 95% confidence intervals are included in brackets. |  |  |  |  |  |  |  |
| *AUC* area under the receiver operating characteristic curve, *ACC* accuracy, *SENS* sensitivity, *SPEC* specificity, *PPV* positive predict value, *NPV* negative predict value. |  |  |  |  |  |  |  |
| *T* training cohort (n = 630), *V* validation cohort (n = 210), *I-T* independent test cohort (n = 218). |  |  |  |  |  |  |  |

**Table 2.** The detailed parameters of VGG16_BN.

| Table 2 The detailed parameters of VGG16_BN. |  |  |  |  |  |  |
| --- | --- | --- | --- | --- | --- | --- |
| Layer name | Input channels | Output channels | Kernel size | Stride | Padding | Output size |
| basic block × 2 | 3 | 64 | 3 | 1 | 1 | [64, 256, 256] |
| max pooling layer |  |  | 2 | 2 | 0 | [64, 128, 128] |
| basic block × 2 | 64 | 128 | 3 | 1 | 1 | [128, 128, 128] |
| max pooling layer |  |  | 2 | 2 | 0 | [128, 64, 64] |
| basic block × 3 | 128 | 256 | 3 | 1 | 1 | [256, 64, 64] |
| max pooling layer |  |  | 2 | 2 | 0 | [256, 32, 32] |
| basic block × 3 | 256 | 512 | 3 | 1 | 1 | [512, 32, 32] |
| max pooling layer |  |  | 2 | 2 | 0 | [512, 16, 16] |
| basic block × 3 | 512 | 512 | 3 | 1 | 1 | [512, 16, 16] |
| max pooling layer |  |  | 2 | 2 | 0 | [512, 8, 8] |
| adaptive average pooling layer |  |  |  |  |  | [512, 7, 7] |
| The basic block was cascade by convolution layer, batch normalization layer, and ReLU. |  |  |  |  |  |  |
| The input size of the model was [3, 256, 256], which followed the format of [channel, height, width]. |  |  |  |  |  |  |

**Table 3.** The performance in prediction of ALN status (N0 vs. N+(1-2)).

| Table 3 The performance in prediction of ALN status (N0 vs. N+(1-2)). |  |  |  |  |  |  |  |
| --- | --- | --- | --- | --- | --- | --- | --- |
| Methods |  | AUC | ACC (%) | SENS (%) | SPEC (%) | PPV (%) | NPV (%) |
| Clinical data only | T | 0.638 [0.595, 0.679] | 61.00 [56.65, 65.23] | 65.62 [56.72, 73.79] | 59.49 [54.43, 64.40] | 34.71 [30.88, 38.75] | 84.06 [80.37, 87.16] |
|  | V | 0.677 [0.602, 0.745] | 74.29 [67.15, 80.58] | 45.45 [30.39, 61.15] | 83.97 [76.55, 89.79] | 48.78 [36.42, 61.29] | 82.09 [77.60, 85.84] |
|  | I-T | 0.627<sup>a, b</sup> [0.551, 0.700] | 72.67 [65.37, 79.18] | 44.74 [28.62, 61.70] | 80.60 [72.88, 86.92] | 39.53 [28.52, 51.73] | 83.72 [79.24, 87.39] |
| DL-CNB model | T | 0.912 [0.884, 0.935] | 82.24 [78.67, 85.44] | 97.66 [93.30, 99.51] | 77.18 [72.69, 81.25] | 58.41 [53.87, 62.81] | 99.01 [97.04, 99.68] |
|  | V | 0.756 [0.685, 0.817] | 59.43 [51.76, 66.77] | 97.73 [87.98, 99.94] | 46.56 [37.81, 55.48] | 38.05 [34.22, 42.04] | 98.39 [89.70, 99.77] |
|  | I-T | 0.845<sup>c</sup> [0.782, 0.895] | 80.23 [73.49, 85.90] | 73.68 [56.90, 86.60] | 82.09 [74.53, 88.17] | 53.85 [43.66, 63.72] | 91.67 [86.53, 94.96] |
| DL-CNB+C model | T | 0.936 [0.911, 0.955] | 84.17 [80.74, 87.21] | 95.31 [90.08, 98.26] | 80.51 [76.23, 84.33] | 61.62 [56.66, 66.34] | 98.12 [95.99, 99.13] |
|  | V | 0.789 [0.721, 0.847] | 66.29 [58.76, 73.24] | 84.09 [69.93, 93.36] | 60.31 [51.39, 68.74] | 41.57 [35.72, 47.67] | 91.86 [84.94, 95.76] |
|  | I-T | 0.878 [0.819, 0.923] | 84.30 [77.99, 89.39] | 71.05 [54.10, 84.58] | 88.06 [81.33, 93.02] | 62.79 [50.52, 73.61] | 91.47 [86.65, 94.66] |
| 95% confidence intervals are included in brackets.<br>*AUC* area under the receiver operating characteristic curve, *ACC* accuracy, *SENS* sensitivity, *SPEC* specificity, *PPV* positive predict value, *NPV* negative predict value.<br>*T* training cohort (n = 518), *V* validation cohort (n = 175), *I-T* independent test cohort (n = 172).<br><sup>a</sup>Indicates $p = 0.0004$, Delong et al. in comparison with DL-CNB model in independent test cohort.<br><sup>b</sup>Indicates $p < 0.0001$, Delong et al. in comparison with DL-CNB+C model in independent test cohort.<br><sup>c</sup>Indicates $p = 0.1148$, Delong et al. in comparison with DL-CNB+C model in independent test cohort. |  |  |  |  |  |  |  |

**Table 4.** The performance in prediction of ALN status (N0 vs. N+(≥3)).

| Table 4 The performance in prediction of ALN status (N0 vs. N+(≥3)). |  |  |  |  |  |  |  |
| --- | --- | --- | --- | --- | --- | --- | --- |
| Methods |  | AUC | ACC (%) | SENS (%) | SPEC (%) | PPV (%) | NPV (%) |
| Clinical data only | T | 0.680 [0.638, 0.721] | 66.67 [62.39, 70.75] | 65.83 [56.62, 74.24] | 66.92 [62.01, 71.58] | 37.98 [33.59, 42.58] | 86.42 [83.10, 89.18] |
|  | V | 0.748 [0.675, 0.813] | 71.52 [63.98, 78.26] | 76.47 [58.83, 89.25] | 70.23 [61.62, 77.90] | 40.00 [32.57, 47.92] | 92.00 [86.13, 95.51] |
|  | I-T | 0.629<sup>a, b</sup> [0.553, 0.701] | 69.36 [61.92, 76.14] | 53.85 [37.18, 69.91] | 73.88 [65.59, 81.08] | 37.50 [28.54, 47.40] | 84.62 [79.43, 88.68] |
| DL-CNB model | T | 0.906 [0.877, 0.930] | 81.57 [77.93, 84.84] | 93.33 [87.29, 97.08] | 77.95 [73.50, 81.97] | 56.57 [51.79, 61.23] | 97.44 [95.10, 98.67] |
|  | V | 0.755 [0.682, 0.819] | 64.24 [56.42, 71.54] | 91.18 [76.32, 98.14] | 57.25 [48.32, 65.85] | 35.63 [30.67, 40.92] | 96.15 [89.36, 98.67] |
|  | I-T | 0.837<sup>c</sup> [0.773, 0.888] | 69.94 [62.52, 76.67] | 92.31 [79.13, 98.38] | 63.43 [54.68, 71.58] | 42.35 [36.61, 48.31] | 96.59 [90.46, 98.83] |
| DL-CNB+C model | T | 0.918 [0.891, 0.940] | 82.16 [78.55, 85.38] | 91.67 [85.21, 95.93] | 79.23 [74.86, 83.15] | 57.59 [52.62, 62.42] | 96.87 [94.45, 98.25] |
|  | V | 0.761 [0.689, 0.824] | 66.06 [58.29, 73.24] | 79.41 [62.10, 91.30] | 62.60 [53.72, 70.89] | 35.53 [29.40, 42.16] | 92.13 [85.66, 95.83] |
|  | I-T | 0.838 [0.774, 0.889] | 71.10 [63.73, 77.73] | 89.74 [75.78, 97.13] | 65.67 [56.98, 73.65] | 43.21 [37.04, 49.60] | 95.65 [89.61, 98.25] |
| 95% confidence intervals are included in brackets.<br>*AUC* area under the receiver operating characteristic curve, *ACC* accuracy, *SENS* sensitivity, *SPEC* specificity, *PPV* positive predict value, *NPV* negative predict value.<br>*T* training cohort (n = 510), *V* validation cohort (n = 165), *I-T* independent test cohort (n = 173).<br><sup>a</sup>Indicates $p = 0.0005$, Delong et al. in comparison with DL-CNB model in independent test cohort.<br><sup>b</sup>Indicates $p < 0.0001$, Delong et al. in comparison with DL-CNB+C model in independent test cohort.<br><sup>c</sup>Indicates $p = 0.9689$, Delong et al. in comparison with DL-CNB+C model in independent test cohort. |  |  |  |  |  |  |  |

**Table 5.** The subgroup performance in prediction of ALN status by DL-CNB+C model (N0 vs. N(+)).

|  |  |  |  |  |  |  |  |  |  |  |
| --- | --- | --- | --- | --- | --- | --- | --- | --- | --- | --- |
| Characteristics | Value |  | AUC | ACC (%) | SENS (%) | SPEC (%) | PPV (%) | NPV (%) |  | *p* |
| Age ≤ 50 | yes | I-T | 0.918 [0.825, 0.971] | 82.09 [70.80, 90.39] | 93.33 [77.93, 99.18] | 72.97 [55.88, 86.21] | 73.68 [62.05, 82.74] | 93.10 [77.72, 98.12] |  | **0.0151** |
|  | no | I-T | 0.794 [0.720, 0.855] | 66.89 [58.77, 74.32] | 90.74 [79.70, 96.92] | 53.61 [43.19, 63.8] | 52.13 [46.38, 57.82] | 91.23 [81.56, 96.07] |  |  |
| T stage | T1 | I-T | 0.833 [0.754, 0.895] | 71.90 [63.01, 79.69] | 89.19 [74.58, 96.97] | 64.29 [53.08, 74.45] | 52.38 [44.70, 59.95] | 93.10 [84.07, 97.19] |  | 0.7426 |
|  | T2 | I-T | 0.814 [0.722, 0.886] | 71.13 [61.05, 79.89] | 93.62 [82.46, 98.66] | 50.00 [35.53, 64.47] | 63.77 [56.91, 70.11] | 89.29 [72.93, 96.27] |  |  |
| ER | positive | I-T | 0.853 [0.789, 0.903] | 82.53 [75.88, 87.98] | 89.23 [79.06, 95.56] | 78.22 [68.90, 85.82] | 72.50 [64.34, 79.39] | 91.86 [84.76, 95.81] |  | 0.1253 |
|  | negative | I-T | 0.737 [0.596, 0.849] | 67.31 [52.89, 79.67] | 73.68 [48.80, 90.85] | 63.64 [45.12, 79.60] | 53.85 [40.83, 66.36] | 80.77 [65.47, 90.30] |  |  |
| PR | positive | I-T | 0.839 [0.772, 0.893] | 72.61 [64.93, 79.42] | 96.61 [88.29, 99.59] | 58.16 [47.77, 68.05] | 58.16 [52.28, 63.82] | 96.61 [87.84, 99.12] |  | 0.6591 |
|  | negative | I-T | 0.811 [0.690, 0.900] | 70.49 [57.43, 81.48] | 80.00 [59.30, 93.17] | 63.89 [46.22, 79.18] | 60.61 [48.85, 71.25] | 82.14 [66.92, 91.27] |  |  |
| HER2 | positive | I-T | 0.800 [0.677, 0.892] | 66.67 [53.31, 78.31] | 88.89 [65.29, 98.62] | 57.14 [40.96, 72.28] | 47.06 [37.68, 56.65] | 92.31 [75.99, 97.85] |  | 0.5238 |
|  | negative | I-T | 0.842 [0.776, 0.895] | 74.05 [66.49, 80.69] | 92.42 [83.20, 97.49] | 60.87 [50.14, 70.88] | 62.89 [56.54, 68.81] | 91.80 [82.60, 96.35] |  |  |
| 95% confidence intervals are included in brackets. |  |  |  |  |  |  |  |  |  |  |
| *AUC* area under the receiver operating characteristic curve, *ACC* accuracy, *SENS* sensitivity, *SPEC* specificity, *PPV* positive predict value, *NPV* negative predict value. |  |  |  |  |  |  |  |  |  |  |
| *I-T* independent test group, *ER* estrogen receptor, *PR* progesterone receptor, *HER-2* human epidermal growth factor receptor-2, *LNM* lymph node metastasis. |  |  |  |  |  |  |  |  |  |  |

**Figure 1.**
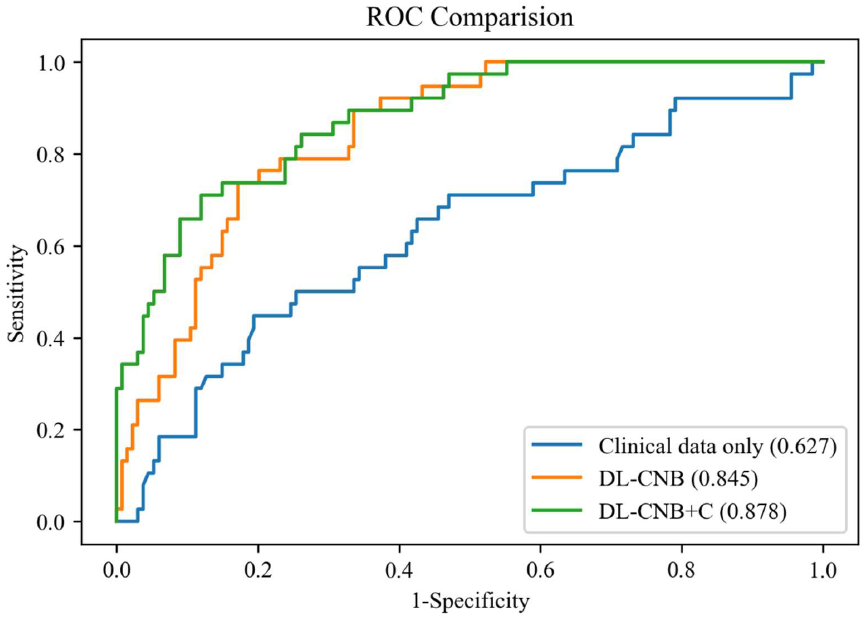
Comparison of receiver operating characteristic (ROC) curves between different models for predicting disease-free axilla (N0) and low metastatic burden of axillary disease (N+(1-2)). Numbers in parentheses are AUCs.

**Figure 2.**
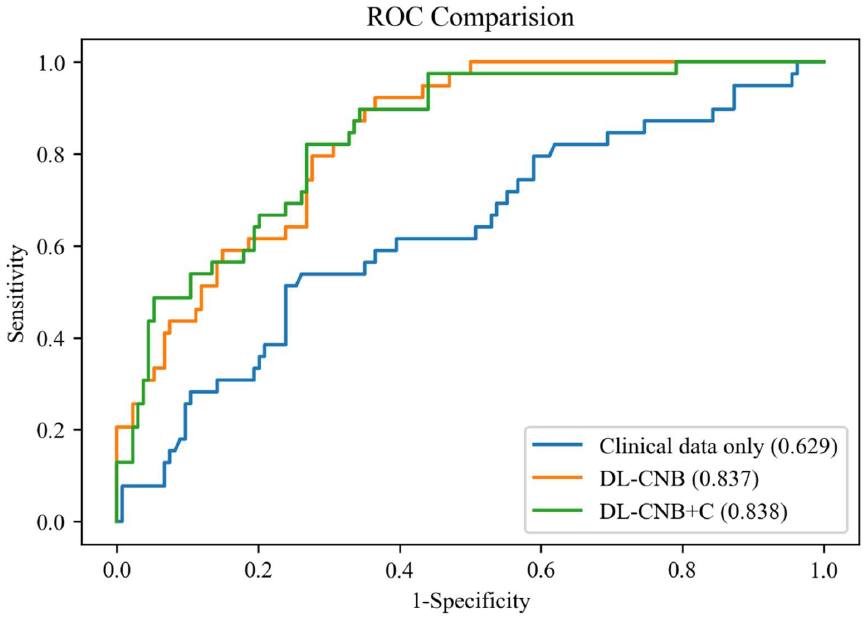
Comparison of receiver operating characteristic (ROC) curves between different models for predicting disease-free axilla (N0) and heavy metastatic burden of axillary disease (N+(≥3)). Numbers in parentheses are AUCs.

## Reference

1. Siegel RL, Miller KD, Jemal A. Cancer statistics, 2019. CA Cancer J Clin (2019) 69(1):7–34. doi: 10.3322/caac.21551.

2. Ahmed M, Purushotham AD, Douek M. Novel techniques for sentinel lymph node biopsy in breast cancer: a systematic review. Lancet Oncol (2014) 15(8):e351–62. doi: 10.1016/S1470-2045(13)70590-4.

3. Kootstra J, Hoekstra-Weebers JEHM, Rietman H, de Vries J, Baas P, Geertzen JHB, et al. Quality of life after sentinel lymph node biopsy or axillary lymph node dissection in stage I/II breast cancer patients: a prospective longitudinal study. Ann Surg Oncol (2008) 15(9):2533–41. doi: 10.1245/s10434-008-9996-9. PubMed Central PMCID: PMCPMC2518082.

4. Wilke LG, McCall LM, Posther KE, Whitworth PW, Reintgen DS, Leitch AM, et al. Surgical complications associated with sentinel lymph node biopsy: results from a prospective international cooperative group trial. Ann Surg Oncol (2006) 13(4):491–500. doi: 10.1245/ASO.2006.05.013.

5. Manca G, Rubello D, Tardelli E, Giammarile F, Mazzarri S, Boni G, et al. Sentinel Lymph Node Biopsy in Breast Cancer: Indications, Contraindications, and Controversies. Clin Nucl Med (2016) 41(2):126–33. doi: 10.1097/RLU.0000000000000985.

6. Hindié E, Groheux D, Brenot-Rossi I, Rubello D, Moretti J-L, Espié M. The sentinel node procedure in breast cancer: nuclear medicine as the starting point. J Nucl Med (2011) 52(3):405–14. doi: 10.2967/jnumed.110.081711.

7. Dihge L, Vallon-Christersson J, Hegardt C, Saal LH, Häkkinen J, Larsson C. et al. Prediction of lymph node metastasis in breast cancer by gene expression and clinicopathological models: development and validation within a population-based cohort. Clin Cancer Res (2019). doi: 10.1158/1078-0432.CCR-19-0075.

8. Shiino S, Matsuzaki J, Shimomura A, Kawauchi J, Takizawa S, Sakamoto H, et al. Serum miRNA–based prediction of axillary lymph node metastasis in breast cancer. Clin Cancer Res (2019). doi: 10.1158/1078-0432.CCR-18-1414.

9. Zheng X, Yao Z, Huang Y, Yu Y, Wang Y, Liu Y, et al. Deep learning radiomics can predict axillary lymph node status in early-stage breast cancer. Nat Commun (2020) 11(1):1236. doi: 10.1038/s41467-020-15027-z. PubMed Central PMCID: PMCPMC7060275.

10. Luo J, Ning Z, Zhang S, Feng Q, Zhang Y. Bag of deep features for preoperative prediction of sentinel lymph node metastasis in breast cancer. Phys Med Biol (2018) 63(24):245014. doi: 10.1088/1361-6560/aaf241.

11. Campanella G, Hanna MG, Geneslaw L, Miraflor A, Werneck Krauss Silva V, Busam KJ, et al. Clinical-grade computational pathology using weakly supervised deep learning on whole slide images.Nat Med (2019) 25(8):1301–9. doi: 10.1038/s41591-019-0508-1. PubMed Central PMCID: PMCPMC7418463.

12. Gu F, Burlutskiy N, Andersson M, Wilén LK. Multi-resolution Networks for Semantic Segmentation in Whole Slide Images. Computational Pathology and Ophthalmic Medical Image Analysis; 2018: Springer International Publishing (2018). p. 11-8. doi: 10.1007/978-3-030-00949-6_2.

13. Mei K, Zhu C, Jiang L, Liu J, Qiao Y. Cross-Stained Segmentation from Renal Biopsy Images Using Multi-Level Adversarial Learning. IEEE International Conference on Acoustics, Speech and Signal Processing; 2020: ieeexplore.ieee.org (2020). p. 1424–8. doi: 10.1109/ICASSP40776.2020.9054505.

14. Jiang L, Chen W, Dong B, Mei K, Zhu C, Liu J, et al. A Deep Learning-Based Approach for Glomeruli Instance Segmentation from Multistained Renal Biopsy Pathologic Images. Am J Pathol (2021) 191(8):1431–41. doi: 10.1016/j.ajpath.2021.05.004.

15. Zhu C, Mei K, Peng T, Luo Y, Liu J, Wang Y, et al. Multi-level colonoscopy malignant tissue detection with adversarial CAC-UNet. Neurocomputing (2021) 438:165–83. doi: 10.1016/j.neucom.2020.04.154.

16. Feng R, Liu X, Chen J, Chen DZ, Gao H, Wu J. A Deep Learning Approach for Colonoscopy Pathology WSI Analysis: Accurate Segmentation and Classification. IEEE J Biomed Health Inform (2020). doi: 10.1109/JBHI.2020.3040269.

17. Iizuka O, Kanavati F, Kato K, Rambeau M, Arihiro K, Tsuneki M. Deep Learning Models for Histopathological Classification of Gastric and Colonic Epithelial Tumours. Sci Rep (2020) 10(1):1504. doi: 10.1038/s41598-020-58467-9. PubMed Central PMCID: PMCPMC6992793.

18. Song Z, Zou S, Zhou W, Huang Y, Shao L, Yuan J, et al. Clinically applicable histopathological diagnosis system for gastric cancer detection using deep learning. Nat Commun (2020) 11(1):4294. doi: 10.1038/s41467-020-18147-8. PubMed Central PMCID: PMCPMC7453200.

19. Hu Y, Su F, Dong K, Wang X, Zhao X, Jiang Y, et al. Deep learning system for lymph node quantification and metastatic cancer identification from whole-slide pathology images. Gastric Cancer (2021) 24(4):868–77. doi: 10.1007/s10120-021-01158-9.

20. Zhao Y, Yang F, Fang Y, Liu H, Zhou N, Zhang J, et al. Predicting lymph node metastasis using histopathological images based on multiple instance learning with deep graph convolution. Proceedings of the IEEE Conference on Computer Vision and Pattern Recognition; 2020: openaccess.thecvf.com (2020). p. 4837–46. doi: 10.1109/CVPR42600.2020.00489.

21. Steiner DF, MacDonald R, Liu Y, Truszkowski P, Hipp JD, Gammage C, et al. Impact of Deep Learning Assistance on the Histopathologic Review of Lymph Nodes for Metastatic Breast Cancer. Am J Surg Pathol (2018) 42(12):1636–46. doi: 10.1097/PAS.0000000000001151. PubMed Central PMCID: PMCPMC6257102.

22. Harmon SA, Sanford TH, Brown GT, Yang C, Mehralivand S, Jacob JM, et al. Multiresolution Application of Artificial Intelligence in Digital Pathology for Prediction of Positive Lymph Nodes From Primary Tumors in Bladder Cancer. JCO Clin Cancer Inform (2020) 4:367–82. doi: 10.1200/CCI.19.00155. PubMed Central PMCID: PMCPMC7259877.

23. Ilse M, Tomczak J, Welling M. Attention-based Deep Multiple Instance Learning. Proceedings of the International Conference on Machine Learning; 2018. p. 2127–36.

24. Das K, Conjeti S, Roy AG, Chatterjee J, Sheet D. Multiple instance learning of deep convolutional neural networks for breast histopathology whole slide classification. IEEE International Symposium on Biomedical Imaging; 2018: ieeexplore.ieee.org (2018). p. 578–81. doi: 10.1109/ISBI.2018.8363642.

25. Sudharshan PJ, Petitjean C, Spanhol F, Oliveira LE, Heutte L, Honeine P. Multiple instance learning for histopathological breast cancer image classification. Expert Syst Appl (2019) 117:103–11. doi: 10.1016/j.eswa.2018.09.049.

26. Couture HD, Marron JS, Perou CM, Troester MA, Niethammer M. Multiple Instance Learning for Heterogeneous Images: Training a CNN for Histopathology. Medical Image Computing and Computer Assisted Intervention; 2018: Springer International Publishing (2018). p. 254–62. doi: 10.1007/978-3-030-00934-2_29.

27. Krizhevsky A, Sutskever I, Hinton GE. Imagenet classification with deep convolutional neural networks. Adv Neural Inf Process Syst (2012) 25:1097–105.

28. Simonyan K, Zisserman A. Very Deep Convolutional Networks for Large-Scale Image Recognition. International Conference on Learning Representations; 2015(2015).

29. He K, Zhang X, Ren S, Sun J. Deep residual learning for image recognition. Proceedings of the IEEE Conference on Computer Vision and Pattern Recognition; 2016: openaccess.thecvf.com (2016). p. 770–8. doi: 10.1109/CVPR.2016.90.

30. Huang G, Liu Z, Van Der Maaten L, Weinberger KQ. Densely connected convolutional networks. Proceedings of the IEEE Conference on Computer Vision and Pattern Recognition; 2017: openaccess.thecvf.com (2017). p. 4700–8. doi: 10.1109/CVPR.2017.243.

31. Szegedy C, Vanhoucke V, Ioffe S, Shlens J, Wojna Z. Rethinking the inception architecture for computer vision. Proceedings of the IEEE Conference on Computer Vision and Pattern Recognition; 2016: openaccess.thecvf.com (2016). p. 2818–26. doi: 10.1109/CVPR.2016.308.

32. Feng J, Zhou Z-H. Deep MIML Network. Proceedings of the AAAI Conference on Artificial Intelligence; 2017: ojs.aaai.org (2017). p. 1884–90.

33. Pinheiro PO, Collobert R. From image-level to pixel-level labeling with convolutional networks. Proceedings of the IEEE Conference on Computer Vision and Pattern Recognition; 2015: openaccess.thecvf.com (2015). p. 1713–21. doi: 10.1109/CVPR.2015.7298780.

34. Zhu W, Lou Q, Vang YS, Xie X. Deep Multi-instance Networks with Sparse Label Assignment for Whole Mammogram Classification. Medical Image Computing and Computer Assisted Intervention; 2017: Springer International Publishing (2017). p. 603–11. doi: 10.1007/978-3-319-66179-7_69.

35. Loshchilov I, Hutter F. SGDR: Stochastic Gradient Descent with Warm Restarts. International Conference on Learning Representations; 2017: OpenReview.net (2017).

36. Mueller JL, Gallagher JE, Chitalia R, Krieger M, Erkanli A, Willett RM, et al. Rapid staining and imaging of subnuclear features to differentiate between malignant and benign breast tissues at a point-of-care setting. J Cancer Res Clin Oncol (2016) 142(7):1475–86. doi: 10.1007/s00432-016-2165-9. PubMed Central PMCID: PMCpmc4900949.

37. Radhakrishnan A, Damodaran K, Soylemezoglu AC, Uhler C, Shivashankar GV. Machine Learning for Nuclear Mechano-Morphometric Biomarkers in Cancer Diagnosis. Sci Rep (2017) 7(1):17946. doi: 10.1038/s41598-017-17858-1. PubMed Central PMCID: PMCPMC5738417.

38. DeLong ER, DeLong DM, Clarke-Pearson DL. Comparing the areas under two or more correlated receiver operating characteristic curves: a nonparametric approach. Biometrics (1988) 44(3):837–45. doi: 10.1038/s41591-019-0508-1.

39. Deng J, Dong W, Socher R, Li L-J, Li K, Fei-Fei L. ImageNet: A large-scale hierarchical image database. Proceedings of the IEEE Conference on Computer Vision and Pattern Recognition; 2009: ieeexplore.ieee.org (2009). p. 248–55.

40. Zhou L-Q, Wu X-L, Huang S-Y, Wu G-G, Ye H-R, Wei Q, et al. Lymph Node Metastasis Prediction from Primary Breast Cancer US Images Using Deep Learning. Radiology (2020) 294(1):19–28. doi: 10.1148/radiol.2019190372.

41. Yang X, Wu L, Ye W, Zhao K, Wang Y, Liu W, et al. Deep Learning Signature Based on Staging CT for Preoperative Prediction of Sentinel Lymph Node Metastasis in Breast Cancer. Acad Radiol (2020) 27(9):1226–33. doi: 10.1016/j.acra.2019.11.007.

42. Calhoun KE, Anderson BO. Needle biopsy for breast cancer diagnosis: a quality metric for breast surgical practice. J Clin Oncol (2014) 32(21):2191–2. doi: 10.1200/JCO.2014.55.6324.

43. Acs B, Rantalainen M, Hartman J. Artificial intelligence as the next step towards precision pathology. J Intern Med (2020) 288(1):62–81. doi: 10.1111/joim.13030.

44. Jaber MI, Song B, Taylor C, Vaske CJ, Benz SC, Rabizadeh S, et al. A deep learning image-based intrinsic molecular subtype classifier of breast tumors reveals tumor heterogeneity that may affect survival. Breast Cancer Res (2020) 22(1):12. doi: 10.1186/s13058-020-1248-3. PubMed Central PMCID: PMCPMC6988279.

45. Whitney J, Corredor G, Janowczyk A, Ganesan S, Doyle S, Tomaszewski J, et al. Quantitative nuclear histomorphometry predicts oncotype DX risk categories for early stage ER+ breast cancer. BMC Cancer (2018) 18(1):610. doi: 10.1186/s12885-018-4448-9. PubMed Central PMCID: PMCPMC5977541.

46. Lu C, Romo-Bucheli D, Wang X, Janowczyk A, Ganesan S, Gilmore H, et al. Nuclear shape and orientation features from H&E images predict survival in early-stage estrogen receptor-positive breast cancers. Lab Invest (2018) 98(11):1438–48. doi: 10.1038/s41374-018-0095-7. PubMed Central PMCID: PMCpmc6214731.

47. Lee G, Veltri RW, Zhu G, Ali S, Epstein JI, Madabhushi A. Nuclear Shape and Architecture in Benign Fields Predict Biochemical Recurrence in Prostate Cancer Patients Following Radical Prostatectomy: Preliminary Findings. European Urology Focus (2017) 3(4):457–66. doi: 10.1016/j.euf.2016.05.009. PubMed Central PMCID: PMCpmc5537035.

